# Myopia and hyperopia are associated with opposite chronotypes in a sample of 71,016 individuals

**DOI:** 10.1101/2024.02.13.24302718

**Authors:** Teele Palumaa, Nele Taba, Maris Teder-Laving, Kadi-Liis Kivi, Kadri Reis, Urmo Võsa, Estonian Biobank Research Team, Tõnu Esko, Erik Abner

**Affiliations:** Estonian Genome Centre, Institute of Genomics, University of Tartu, Tartu, Estonia; Department of Ophthalmology, Emory University, Atlanta, USA; East Tallinn Central Hospital, Eye Clinic, Tallinn, Estonia

## Abstract

Myopia, projected to affect half of the global population by 2050, is a growing healthcare concern. Chronotype, an output of the human biological clock, and sleep parameters have been associated with several diseases, including myopia. We explored the connection between refractive errors and sleep and circadian rhythm parameters by employing a sample of 71,016 adults who completed the Munich Chronotype Questionnaire in the Estonian Biobank. After accounting for possible confounders, such as age, sex, education level, and duration of daylight exposure, we observed that individuals with late chronotype, characterised by a delayed sleep-wake pattern on free days, had higher odds for myopia. In contrast, early chronotype was associated with hyperopia. Furthermore, increased social jet lag and reduced sleep duration were associated with both myopia and hyperopia. These results emphasise the complex interplay between circadian rhythms and sleep in refractive development, with potential implications for public health and clinical practice.

## Introduction

Myopia, or short-sightedness, is an ocular condition characterised by impaired distant vision. It has increasingly been recognised as a substantial global public health concern as its prevalence is rising rapidly and is estimated to reach 50% by 2050^1^. During eye development, the length of the eye and the structures that bend incoming light rays, the cornea, and the crystalline lens, must become balanced to focus light on the retina, where visual signalling is initiated. Myopia results from a failure in this refractive development process, caused primarily by excessive eyeball elongation, leading to light entering the eye and focusing in front of the retina. The opposite condition, where the eye remains too short, is termed hyperopia or long-sightedness. Although refractive development occurs during childhood and is complete by early adulthood, the shorter axial length in hyperopia is initially compensated by an elastic crystalline lens^2^. Thus, the hyperopes remain asymptomatic and do not receive the diagnosis until later in adulthood^2^. Although myopia and hyperopia can be corrected with spectacles or contact lenses, the mechanical strain induced by the excessive elongation of the eye in myopic individuals increases the susceptibility to severe ocular pathologies that could jeopardise vision in later stages of life. These include myopic macular degeneration, retinal detachment, glaucoma, and cataracts^3^. This not only impairs the quality of life of affected individuals but also imposes substantial social and economic costs^4^. Given the considerable impact of myopia, a better understanding of the mechanisms governing its development and associated risk factors is required.

Several studies have suggested a connection between refractive development and circadian rhythms. For example, the axial length of the eye displays rhythmicity over a day in humans^5^ as well as in chickens^6^. Furthermore, the rhythmicity of axial length is disrupted in chickens developing myopia^7^. Additionally, a meta-analysis of genome-wide association studies (GWAS), which included over 500,000 study subjects, revealed that genes associated with myopia were enriched for genes regulating circadian rhythms^8^. These findings have led to the analysis of behavioural circadian rhythms in myopia. Chronotype, an individual’s inherent predilection for sleep-wake timing, is dictated by the individual’s biological clock^9^. This preference spans a spectrum from early chronotypes, or “larks” (individuals with a propensity for early rising and retiring), to late chronotypes, or “owls” (individuals who favour delayed sleep and wake times)^9^. The establishment of activity patterns is a result of the extensive interaction between the biological clock, the natural cycle of light and darkness, as well as social schedules, often referred to as the social clock^9^. Misalignment of the biological and social clock, termed social jet lag (SJL)^10^, is associated with various adverse health outcomes, including metabolic and cardiovascular diseases^11^. SJL is strongly predicted by chronotype^12^ as later chronotype is associated with increased SJL. Furthermore, chronotype is intricately linked to other sleep parameters, as late chronotype has been associated with worse sleep quality^13–15^ and shorter sleep duration^13^. Collectively, the parameters pertaining to sleep and circadian rhythms are complexly interconnected, thereby posing challenges to their investigation.

Various sleep parameters, particularly bedtime, sleep quality, and duration, have been investigated in myopia but, thus far, have yielded mixed results. Several studies have found that children with myopia sleep later than children without myopia^16–19^, while some have found no association^20–23^. Myopia has been associated with lower sleep quality in some studies^16,24^, while no associations have been found in others^25^. Myopia has been associated with shorter sleep duration^16,26^, while other investigations have not confirmed this finding^17,20–25^. Chronotype has also been studied in the context of myopia, but no definitive associations have been established. Some studies have reported an association between myopia and late chronotype^16,27,28^, while in other reports, the circadian rhythms of myopes and non-myopes did not differ^29^ and self-reporting as “morning” or “evening” type was not associated with myopia progression^18^. In conclusion, the existing research presents a mosaic of findings suggesting potentially complex relationships between circadian rhythms and refractive errors.

Leveraging data from the Estonian Biobank (EstBB), we adopted an exhaustive methodology to explore the intricate relationships among refractive errors, chronotype, and sleep parameters. We first characterised chronotype and sleep parameters obtained with the Munich Chronotype Questionnaire (MCTQ)^9^ in the EstBB. Using logistic regression, we then investigated the associations between chronotype, SJL and sleep duration with myopia and hyperopia in ∼71,000 individuals aged between 18 and 70 years. We found that late chronotype was associated with myopia, while early chronotype was linked with hyperopia. Increased SJL and shorter sleep duration were associated with increased odds for both conditions. These associations remained significant after accounting for factors shown to be associated with myopia in previous studies, such as time spent in daylight^30–33^, the highest level of education, birth season^34,35^ and photoperiod at birth^34^, and the first ten genetic principal components.

## Results

### Characterisation of chronotype and sleep parameters in the Estonian Biobank

The MCTQ^9^ includes questions about daily sleep habits, separately for work and work-free days, and was filled out by 136,618 EstBB participants. MCTQ defines chronotype as the midpoint of sleep on work-free days adjusted for sleep debt accumulated over workdays (midpoint of sleep on free days, sleep debt corrected, MSFsc). With MCTQ, chronotype can only be accurately estimated for those who have not done any shift work over the previous three months and who wake up without an alarm clock on free days^9^. Individuals not fulfilling these criteria were not included in the analyses presented below (see Supplementary Fig. 1 for sample selection workflow). SJL is defined as the absolute difference between the midpoint of sleep on free days and the midpoint of sleep on workdays^9^. Weekly average sleep duration was calculated based on the sleep duration on working days and free days^9^.

In our study sample, chronotype could be calculated for 79,247 participants aged between 18 and 80 years. We observed significant variability in chronotype and sleep parameters across age groups between sexes (Fig. 1a-c, Table 1). In line with previous reports^12,36^, chronotype was significantly later in young adults, and it became gradually earlier with increasing age. Males exhibited later chronotypes until the mid-40s and became earlier chronotypes from the 50s, similar to what has been documented previously^12,36^. As shown before^10,12^, SJL decreased with age and became negligible by mid-60s. As late chronotype is associated with increased SJL, one would expect to see similar sex differences in chronotype and SJL. Instead, we found that SJL was not significantly different between males and females in early adulthood but was larger in females from 30s to 60s. Consistent with previous reports^37–39^, the average sleep duration decreased with age. Interestingly, we detected a subsequent increase in sleep duration from the mid-60s, which coincides with the statutory retirement age in Estonia. Females slept longer than males throughout early adulthood to the mid-50s, driven primarily by longer sleep on weekends (Supplementary Fig. 2c). An additional determinant of an individual’s sleep patterns is the duration of daylight exposure as an increased daylight exposure has been associated with an earlier chronotype^40^. We found that males reported spending significantly more time in daylight across all ages (Fig. 1d), suggesting that other factors, not daylight exposure, may account for chronotype differences between males and females. Other sleep-related variables of MCTQ are reported in Supplementary Fig. 2.

**Figure 1.**
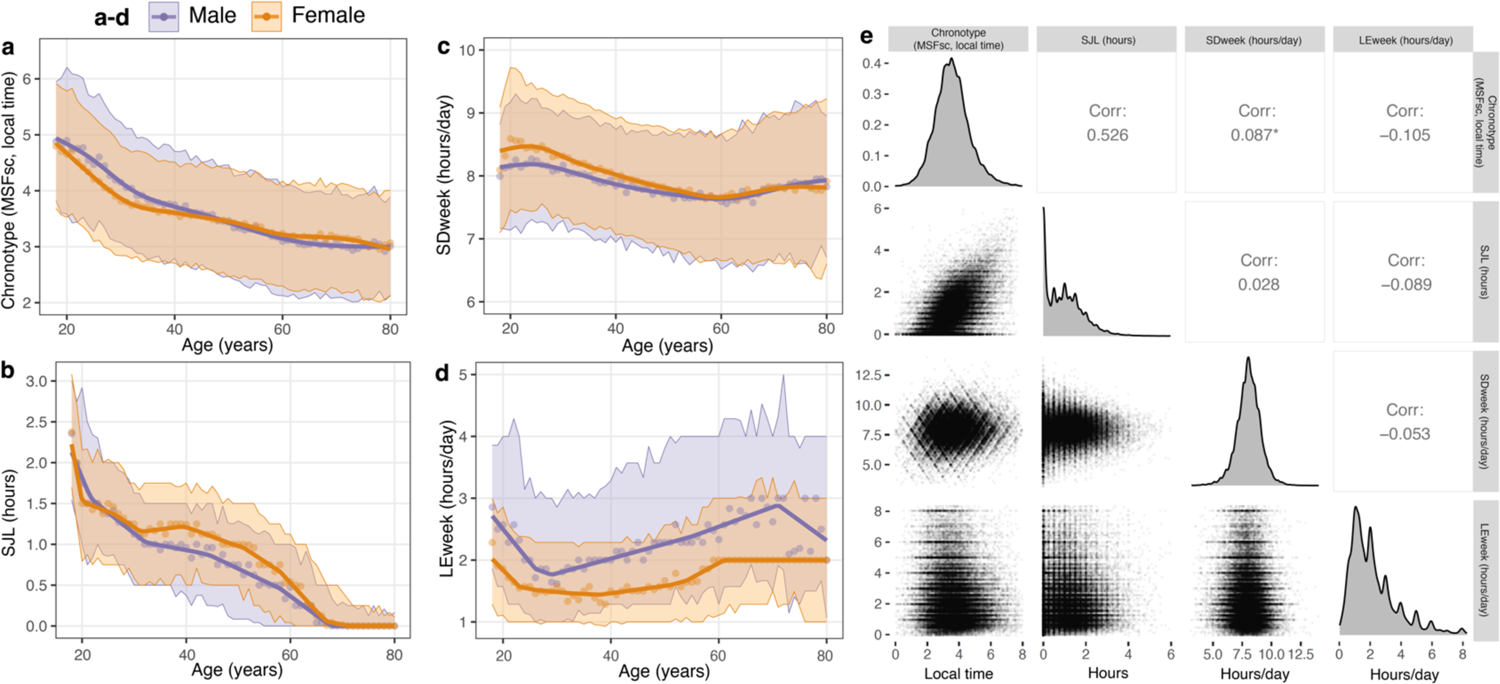
Characterisation of sleep and daylight exposure parameters by age and sex. **a-d** Chronotype (MSFsc, local time) (**a**), social jet lag (SJL) (**b**), weekly average sleep duration per day (SDweek) (**c**), and daily average daylight exposure (LEweek) (**d**) across ages in one-year bins in males and females. **a, c** Points display means for sex and age, highlighted areas indicate ±SD and means are fitted with a generalised additive model. **b, d** Points display medians for sex and age, highlighted areas indicate the interquartile range and medians are fitted with quantile regression. **e** Pairwise correlations between the parameters in **a-d**, reported as Spearman correlation, except * Pearson correlation. Only participants who woke up without an alarm clock on free days and had not done shift work three months prior to replying to the questionnaire were included.

**Table 1.** Characterisation of chronotype, social jet lag and sleep duration by sex and age group in a population aged between 18 and 80 years.

|  | Total N = 79,247 |  | MSFsc, EMM (95% CI)<sup>1</sup> |  |  | SJL, median (95% CI)<sup>2</sup> |  |  | SDweek, EMM (95% CI)<sup>1</sup> |  |  |
| --- | --- | --- | --- | --- | --- | --- | --- | --- | --- | --- | --- |
| Age group (y) | Male | Female | Male | Female | p value | Male | Female | p value | Male | Female | p value |
| 18–25 | 3,053 | 5,199 | 4.73 (4.70–4.77) | 4.51 (4.49–4.54) | **3.5E-22** | 1.54 (1.42–1.58) | 1.50 (1.49–1.50) | 0.34 | 8.18 (8.15–8.22) | 8.46 (8.43–8.48) | **5.6E-33** |
| 26–30 | 2,493 | 4,566 | 4.32 (4.28–4.36) | 3.98 (3.95–4.01) | **2.3E-42** | 1.25 (1.25–1.27) | 1.25 (1.17–1.25) | 0.44 | 8.13 (8.09–8.17) | 8.36 (8.33–8.39) | **7.1E-20** |
| 31–35 | 2,564 | 4,771 | 3.96 (3.92–4.00) | 3.75 (3.72–3.77) | **1.1E-18** | 1.00 (1.00–1.00) | 1.17 (1.13–1.21) | **7.4E-05** | 8.03 (7.99–8.07) | 8.21 (8.18–8.24) | **4.5E-14** |
| 36–40 | 2,746 | 5,299 | 3.78 (3.74–3.81) | 3.63 (3.61–3.66) | **9.8E-10** | 1.00 (1.00–1.08) | 1.21 (1.17–1.25) | **7.0E-05** | 7.90 (7.86–7.93) | 8.06 (8.04–8.09) | **8.2E-13** |
| 41–45 | 2,650 | 5,343 | 3.64 (3.60–3.67) | 3.56 (3.53–3.58) | **0.0008** | 0.92 (0.84–1.00) | 1.13 (1.04–1.13) | **0.0002** | 7.82 (7.78–7.86) | 7.94 (7.92–7.97) | **1.10E-07** |
| 46–50 | 2,733 | 6,043 | 3.50 (3.47–3.54) | 3.48 (3.46–3.51) | 0.31 | 0.75 (0.75–0.75) | 1.00 (1.00–1.00) | **3.3E-11** | 7.74 (7.71–7.78) | 7.84 (7.81–7.86) | **5.30E-05** |
| 51–55 | 2,419 | 5,247 | 3.33 (3.29–3.37) | 3.39 (3.36–3.41) | **0.016** | 0.58 (0.50–0.67) | 0.92 (0.88–0.96) | **3.1E-07** | 7.67 (7.63–7.71) | 7.75 (7.72–7.78) | **0.0012** |
| 56–60 | 2,333 | 4,883 | 3.20 (3.16–3.24) | 3.26 (3.24–3.29) | **0.009** | 0.50 (0.50–0.50) | 0.67 (0.58–0.71) | **0.0004** | 7.66 (7.62–7.70) | 7.66 (7.63–7.69) | 0.93 |
| 61–65 | 1,888 | 4,034 | 3.07 (3.03–3.12) | 3.18 (3.14–3.21) | **0.0002** | 0.25 (0.25–0.29) | 0.25 (0.13–0.25) | 0.49 | 7.64 (7.60–7.69) | 7.70 (7.67–7.73) | **0.046** |
| 66–70 | 1,784 | 3,582 | 3.05 (3.01–3.10) | 3.18 (3.14–3.21) | **1.80E-05** | 0.00 (0.00–0.00) | 0.04 (0.04–0.04) | 0.06 | 7.73 (7.69–7.78) | 7.77 (7.74–7.80) | 0.23 |
| 71–75 | 1,173 | 2,412 | 2.98 (2.93–3.04) | 3.13 (3.09–3.17) | **4.20E-05** | 0.00* | 0.00* | 0.34 | 7.85 (7.80–7.91) | 7.81 (7.77–7.85) | 0.25 |
| 76–80 | 686 | 1,346 | 3.01 (2.94–3.09) | 3 (2.95–3.05) | 0.78 | 0.00* | 0.00* | 0.34 | 7.89 (7.81–7.96) | 7.82 (7.77–7.88) | 0.18 |

<sup>1</sup> Generalised linear model was fitted with two independent variables, age group and sex, along with their interaction, and the dependent variable, MSFsc or SDweek. Post-hoc comparisons were conducted using the emmeans R package. Estimated marginal means (EMMs) and their 95% CI were calculated for each combination of sex and age group. The significance of differences between sexes within each age group was assessed using two-tailed t-tests and the Bonferroni method to adjust for multiple comparisons.
<sup>2</sup> Quantile regression model was fitted with two independent variables, age group and sex, along with their interaction, and the dependent variable SJL. Differences in median values between the sexes within each age group were assessed using the p values associated with the interaction terms between sex and each age group in the quantile regression model. p values < 0.05 are indicated in bold. Medians and 95% CI for medians were estimated using bootstrapping with the boot R package.
CI – confidence interval, EMM – estimated marginal means, MSFsc – mid-point of sleep on free days adjusted for sleep debt, SD – standard deviation, SDweek – weekly average sleep duration, SJL – social jet lag.

To understand to what extent chronotype, SJL, sleep duration, and daily light exposure relate to each other, we analysed the correlation between these variables. We detected a moderate positive correlation between chronotype and SJL (Spearman r_s_ = 0.526) but no correlation between the other variables (Fig. 1e).

### Association between sleep and circadian rhythm parameters with refractive errors

Next, we wanted to analyse the associations between chronotype and sleep parameters with refractive errors. As myopia and hyperopia are opposite conditions—myopia is associated primarily with an increased ocular axial length and hyperopia with a reduced axial length^2^, this approach allows us to investigate factors predisposing eyes to abnormal growth and the factors affecting the directionality of this eye growth. We also included in our analysis factors known to be associated with myopia, including time spent in daylight^30–33^, the highest level of education, birth season^34,35^ and photoperiod at birth^34^. We only included participants aged ≤70 as age-related diseases, e.g. ocular and neurological conditions and eye diseases, can influence one’s sleep patterns and circadian rhythms^41,42^. The workflow for obtaining the sample is presented in Supplementary Figure 1, and descriptive statistics for the obtained sample and refractive error subgroups are presented in Table 2.

**Table 2.** Characteristics of the study population included in the logistic regression analysis.

| Characteristic | Refractive error |  |  |  | p value<sup>1</sup> |
| --- | --- | --- | --- | --- | --- |
|  | Overall<br>N = 71,016 (100%) | None<br>N = 41,832 (58.9%) | Myopia<br>N = 18,516 (26.1%) | Hyperopia<br>N = 10,668 (15.0%) |  |
| Age, median (IQR) | 44 (32, 55) | 43 (32, 53) | 40 (29, 51) | 56 (47, 63) | **<0.001** |
| Sex, n (%) |  |  |  |  | **<0.001** |
| Male | 24,001 (33.8%) | 16,415 (39.2%) | 4,668 (25.2%) | 2,918 (27.4%) |  |
| Female | 47,015 (66.2%) | 25,417 (60.8%) | 13,848 (74.8%) | 7,750 (72.6%) |  |
| Highest level of education, n (%) |  |  |  |  | **<0.001** |
| Basic | 5,477 (7.7%) | 3,531 (8.4%) | 960 (5.2%) | 986 (9.2%) |  |
| Middle | 33,174 (46.7%) | 19,719 (47.1%) | 7,572 (40.9%) | 5,883 (55.1%) |  |
| Higher | 32,365 (45.6%) | 18,582 (44.4%) | 9,984 (53.9%) | 3,799 (35.6%) |  |
| Birth season, n (%) |  |  |  |  | 0.49 |
| Autumn | 16,913 (23.8%) | 9,949 (23.8%) | 4,422 (23.9%) | 2,542 (23.8%) |  |
| Spring | 18,588 (26.2%) | 10,946 (26.2%) | 4,832 (26.1%) | 2,810 (26.3%) |  |
| Summer | 18,505 (26.1%) | 10,806 (25.8%) | 4,879 (26.4%) | 2,820 (26.4%) |  |
| Winter | 17,010 (24.0%) | 10,131 (24.2%) | 4,383 (23.7%) | 2,496 (23.4%) |  |
| Photoperiod at birth (hh:mm), n (%) |  |  |  |  | 0.15 |
| Shortest (06:03-08:23) | 16,472 (23.2%) | 9,768 (23.4%) | 4,325 (23.4%) | 2,379 (22.3%) |  |
| Short (08:25-12:21) | 17,557 (24.7%) | 10,382 (24.8%) | 4,494 (24.3%) | 2,681 (25.1%) |  |
| Long (12:23-16:18) | 18,169 (25.6%) | 10,704 (25.6%) | 4,720 (25.5%) | 2,745 (25.7%) |  |
| Longest (16:18-18:39) | 18,818 (26.5%) | 10,978 (26.2%) | 4,977 (26.9%) | 2,863 (26.8%) |  |
| Ancestry, n (%) |  |  |  |  | **<0.001** |
| Eastern European | 66,483 (93.7%) | 39,142 (93.6%) | 17,485 (94.6%) | 9,856 (92.5%) |  |
| Finnish | 4,239 (6.0%) | 2,516 (6.0%) | 956 (5.2%) | 767 (7.2%) |  |
| Other | 223 (0.3%) | 139 (0.3%) | 51 (0.3%) | 33 (0.3%) |  |
| Unknown | 71 | 35 | 24 | 12 |  |
| Chronotype (MSFsc, local time), mean (SD) | 3.65 (1.09) | 3.66 (1.08) | 3.80 (1.09) | 3.35 (1.04) | **<0.001** |
| SJL (hours), median (IQR) | 1.00 (0.25, 1.50) | 1.00 (0.25, 1.50) | 1.00 (0.50, 1.71) | 0.63 (0.00, 1.25) | **<0.001** |
| SDweek<sup>2</sup> (hours/day), mean (SD) | 7.95 (1.01) | 7.97 (1.00) | 8.00 (0.99) | 7.76 (1.06) | **<0.001** |
| LEweek<sup>3</sup> (hours/day), median (IQR) | 111 (60, 180) | 111 (60, 180) | 94 (60, 154) | 120 (77, 194) | **<0.001** |

<sup>1</sup> To assess the differences in characteristics between the three refractive error groups, the Kruskal-Wallis rank sum test was used for age, MSFsc, SJL, SDweek and LEweek. For all other variables, Pearson's Chi-squared test was used. p values < 0.05 are highlighted in bold.
<sup>2</sup> Average daily sleep duration across a week
<sup>3</sup> Average daily light exposure across a week
LEweek – average daily exposure to daylight, MSFsc – mid-point of sleep on free days adjusted for sleep debt, SDweek – weekly average sleep duration, SJL – social jet lag, IQR – interquartile range, SD – standard deviation.

Myopia frequency was highest in the youngest age group, reaching 37% in the 18-25-year-olds, and decreased with age; conversely, the frequency of hyperopia increased with age (Fig. 2a). The distribution of myopia and hyperopia across ages follows a similar pattern as the estimated prevalence of these refractive errors in Europe^43^. While a total of 12% of males had hyperopia and 19% of males had myopia, the proportion of females with these refractive errors was higher—16% and 29%, respectively (Fig. 2b). Of all myopes, 54% had a higher education degree, this figure was 44% amongst those with no refractive error and 36% in hyperopes (Fig. 2c). The opposite trend was evident for basic and middle education, which were more prevalent in hyperopes and people with no refractive error (Fig. 2c). The distribution of birth season and photoperiod at birth was uniform in myopes, hyperopes and those without a refractive error (Table 2). There were significant differences in chronotype, SJL, average sleep duration and average daily daylight exposure in myopes, hyperopes and individuals with no refractive error across ages (Fig. 2d-g, Table 2).

**Figure 2.**
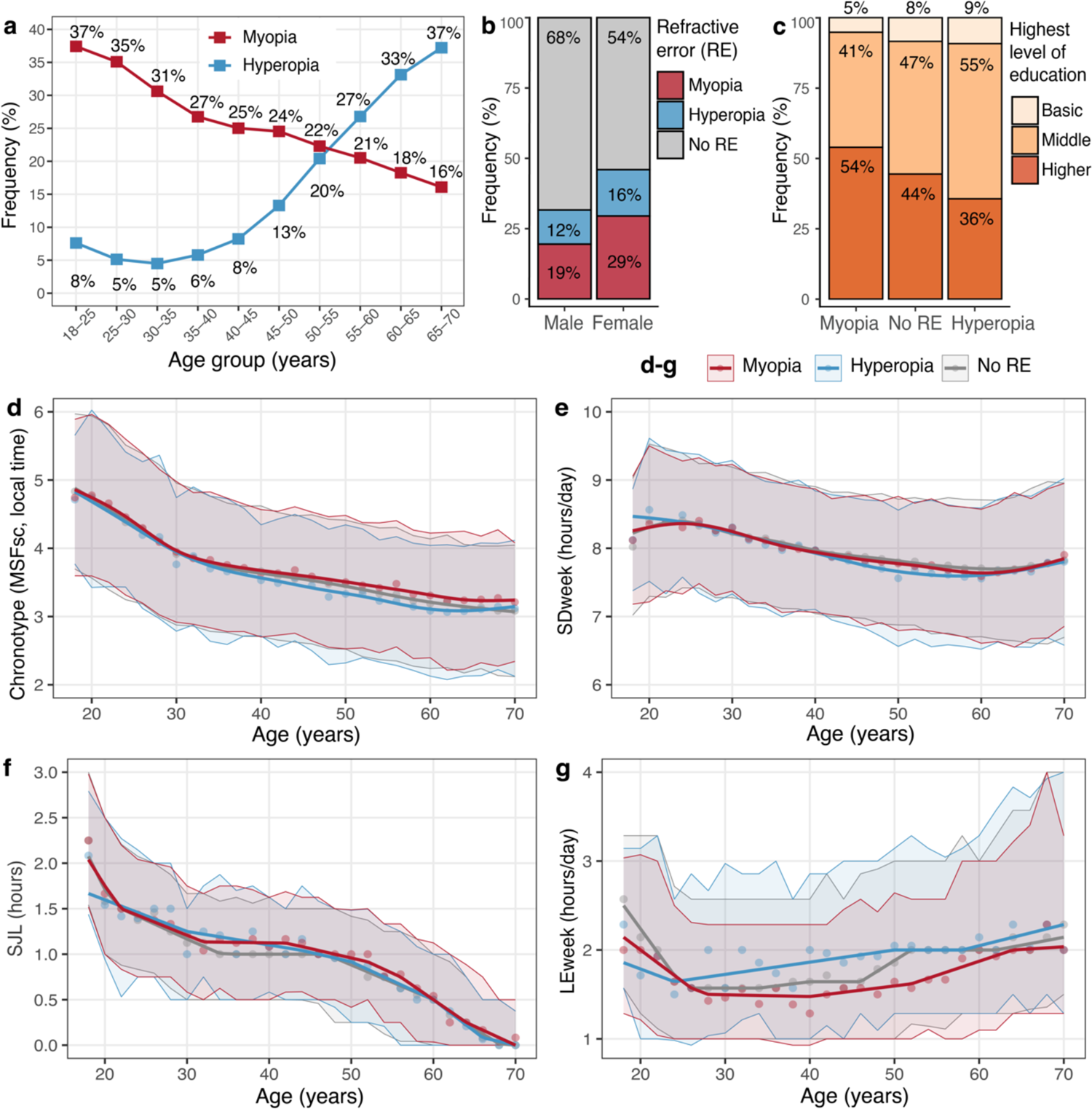
Characterisation of variables significantly associated with myopia and hyperopia in the logistic regression analysis. **a** Frequency of myopia and hyperopia in the study sample based on age at replying to the Munich Chronotype Questionnaire. **b** Frequency of myopia and hyperopia in male and female participants. **c** Distribution of highest level of education by refractive error status. **d**-**g** Chronotype (MSFsc, local time) (**d**), weekly average sleep duration (SDweek) (**e**), social jet lag (SJL) (**f**), and average daily exposure to daylight (LEweek) (**g**) in myopes, hyperopes and participants with no refractive error (RE) across age summarised in 2-year bins. **d, e** points display means for each refractive error status and age group, highlighted areas indicate ±SD and means are fitted with a generalised additive model. **b, d** Points display medians for each refractive error status and age group, highlighted areas indicate the interquartile range and medians are fitted with quantile regression

Next, we used logistic regression analysis to determine whether circadian rhythm and sleep parameters were associated with refractive errors. We first conducted adjusted univariate models to examine the individual effects of each independent variable on the dependent variables myopia and hyperopia while controlling for age, age squared, and sex (Supplementary Table 1). All three sleep and circadian rhythm parameters, chronotype, SJL and sleep duration, were associated with both myopia and hyperopia. Additionally, education level and time spent in daylight showed significant associations with both myopia and hyperopia, and the autumn birth season and longest photoperiod were associated with higher odds of hyperopia compared to the winter birth season and shortest photoperiod.

To further assess whether the detected associations are independent of each other, we employed multivariate models that incorporated all these factors. As refractive errors have a considerable genetic predisposition^8^, we also included in our analysis the first ten genetic principal components calculated from the genetic data of the EstBB participants. The relationships between all studied sleep and circadian parameters remained significant and with similar directionality as in the adjusted univariate analyses (Table 3). Chronotype exhibited an opposite association with refractive errors—one hour later chronotype increased the odds for myopia (OR 1.03, 95% CI 1.01–1.05) and decreased odds for hyperopia (OR 0.95, 95% 0.93–0.98). SJL and sleep duration influenced the odds of myopia and hyperopia in a similar direction. Larger SJL increased the odds for both myopia and hyperopia (per one hour of larger SJL: OR 1.04, 95% CI 1.02–1.07 for myopia; OR 1.09, 95% CI 1.05–1.13 for hyperopia). Shorter sleep was associated with both myopia and hyperopia (per one hour longer sleep duration per day: OR 0.95, 95% CI 0.93–0.97 for myopia, OR 0.92, 95% CI 0.90–0.94 for hyperopia). As expected, more time in daylight decreased the odds for myopia (OR 0.95, 95% CI 0.93–0.97 per additional hour/day). Interestingly, the opposite was evident for hyperopia, more time outdoors was associated with higher odds for hyperopia (OR 1.06, 95% CI 1.04–1.07 per additional hour/day). As has been shown previously, a higher education level increased myopia odds (higher education compared to middle education, OR 1.42, 95% CI 1.36–1.47). Again, the opposite was detected for hyperopia, where higher education was associated with decreased odds (OR 0.73, 95% CI 0.70–0.77). Birth season and photoperiod at birth were not significantly associated with either myopia or hyperopia in the multivariate models.

**Table 3.** Association of myopia and hyperopia with sleep and circadian rhythm parameters.

| Predictors | Myopia |  |  | Hyperopia |  |  |
| --- | --- | --- | --- | --- | --- | --- |
|  | Odds Ratio | 95% CI | p value | Odds Ratio | 95% CI | p value |
| Chronotype (MSFsc, per hour) | 1.03 | 1.01 – 1.05 | **0.002** | 0.95 | 0.93 – 0.98 | **<0.001** |
| SJL (per hour) | 1.04 | 1.02 – 1.07 | **0.001** | 1.09 | 1.05 – 1.13 | **<0.001** |
| SDweek (per hour/day) | 0.95 | 0.93 – 0.97 | **<0.001** | 0.92 | 0.90 – 0.94 | **<0.001** |
| LEweek (per hour/day) | 0.96 | 0.95 – 0.97 | **<0.001** | 1.06 | 1.04 – 1.07 | **<0.001** |
| Education |  |  |  |  |  |  |
| Middle | *Reference* |  |  | *Reference* |  |  |
| Basic | 0.60 | 0.56 – 0.65 | **<0.001** | 1.00 | 0.92 – 1.09 | 0.97 |
| Higher | 1.42 | 1.36 – 1.47 | **<0.001** | 0.73 | 0.70 – 0.77 | **<0.001** |
| Birth season |  |  |  |  |  |  |
| Winter | *Reference* |  |  | *Reference* |  |  |
| Spring | 1.04 | 0.96 – 1.12 | 0.30 | 1.05 | 0.96 – 1.16 | 0.29 |
| Summer | 1.06 | 0.96 – 1.15 | 0.24 | 1.07 | 0.95 – 1.20 | 0.26 |
| Autumn | 1.03 | 0.97 – 1.09 | 0.32 | 1.03 | 0.96 – 1.11 | 0.39 |
| Photoperiod at birth (hh:mm) |  |  |  |  |  |  |
| Shortest (06:03-08:23) | *Reference* |  |  | *Reference* |  |  |
| Short (08:25-12:21) | 0.96 | 0.91 – 1.02 | 0.16 | 1.04 | 0.97 – 1.12 | 0.28 |
| Long (12:23-16:18) | 0.96 | 0.89 – 1.04 | 0.32 | 1.00 | 0.90 – 1.10 | 0.94 |
| Longest (16:18-18:39) | 0.98 | 0.90 – 1.07 | 0.70 | 1.02 | 0.91 – 1.15 | 0.71 |
| Sex, reference: male | 1.83 | 1.76 – 1.91 | **<0.001** | 1.81 | 1.72 – 1.90 | **<0.001** |
| Age | 0.91 | 0.90 – 0.92 | **<0.001** | 0.96 | 0.95 – 0.97 | **<0.001** |
| Age squared / 1,000 | 2.53 | 2.29 – 2.80 | **<0.001** | 2.80 | 2.46 – 3.19 | **<0.001** |
| Intercept | 3.19 | 2.40 – 4.25 | **<0.001** | 0.18 | 0.12 – 0.26 | **<0.001** |
| Observations | 60,348 |  |  | 52,500 |  |  |

Odds ratios were obtained from two multivariable logistic regression models with myopia or hyperopia status as the dependent variable, including participants classified as myopes or hyperopes as cases, respectively, and participants with no refractive error as controls. Both models used MSFsc, SJL, SDweek, LEweek, education, birth season, and photoperiod at birth as predictors, and were adjusted for age, age squared, sex and first ten genetic principal components. *p* values < 0.05 are highlighted in bold.
LEweek –average daily exposure to daylight, MSFsc – mid-point of sleep on free days adjusted for sleep debt, SDweek – weekly average sleep duration, SJL – social jet lag, CI – confidence interval.

For sensitivity analysis, we applied the multivariate model to a sample including participants aged >45 years only. We chose this age as hyperopia often goes undiagnosed until around that age, as the elastic lens compensates for the shorter ocular axial length^2^. We found that the associations with chronotype, SJL, sleep duration, time spent in daylight, and education remained significant with the same directionality (Supplementary Table 2). Collectively, these findings suggest that processes related to SJL and sleep duration are associated with refractive errors in general. In contrast, chronotype is associated with the directionality of refractive error development, revealing a complex relationship between circadian rhythms and sleep and refractive errors.

## Discussion

In this study, we analysed the associations between refractive errors and circadian rhythm and sleep parameters using an extensive chronotype dataset encompassing 71,016 participants from the EstBB. The large age span of our sample enabled us to investigate opposite refractive errors and thus to dissect factors predisposing to eye growth abnormalities in general and factors influencing the directionality of eye growth. One of the most striking findings is the robust bidirectional association of chronotype with refractive errors, where individuals with a later chronotype showed increased odds of having myopia, aligning with previous reports employing smaller samples^22,27,28^. In contrast, an earlier chronotype was linked to a higher likelihood of hyperopia. It should be noted, however, that refractive development occurs primarily in childhood and adolescence^2^, while chronotype was assessed during adulthood in our study sample. Nevertheless, chronotype has a considerable genetic basis, reaching a heritability of 12-20%^44–46^, with some of the most significant genetic hits being genes regulating the circadian clock on a molecular level^44–46^, such as period 1, 2 and 3 (*PER1*, *PER2*, *PER3*), cryptochrome 1 (*CRY1*) and *ARNTL*. In twin studies, self-reported morningness-eveningness has been shown to reach an inheritance of 44% in adolescent twins and 47% in twins in their mid-40s^47^. Furthermore, variations in molecular circadian clock genes underlie extreme sleep-wake cycle phenotypes, for example, the familial advanced sleep-phase syndrome, characterised by a persistent 3-4-hour advanced sleep schedule compared to desired times^48^, is caused by mutations in two core molecular clock genes, *PER2*^49^ and casein kinase 1 delta (*CSNK1D*)^50^. Collectively, these data suggest that chronotype reflects an intrinsic property rather than a transient state of an individual’s sleep-wake cycle. We also found that time spent in daylight as an adult was associated with myopia and hyperopia. We hypothesise that although the amount of time spent outdoors in adulthood would not affect refractive development, it may reflect habits ingrained from childhood.

This association between chronotype and refractive errors raises multiple hypotheses regarding the biological mechanisms influencing eye growth. First, chronotype may influence the development of refractive errors by predisposing individuals to a factor associated with myopia and hyperopia. For example, late chronotypes might spend less time outdoors, as shown in previous reports^9,51^, which is a well-characterised risk factor for myopia^31–33^. Speaking against this hypothesis, we found no correlation between chronotype and time spent outdoors in our study sample (Fig. 1e), and we adjusted our analysis for the reported time spent outdoors. However, these might not have reflected the variables in childhood, as discussed above. Second, refractive errors may influence the development of chronotype—it could be that having myopia or hyperopia shifts individuals’ sleep-wake cycles later and earlier, respectively. The primary brain centres regulating circadian activity are the suprachiasmatic nuclei (SCN), which receive extensive direct input on light levels from the retinas of the eyes^52^. It is known that some aspects of retinal signalling are altered in myopia^53^, which could potentially modify the light input reaching the SCN and result in variations in chronotype. Third, the link between chronotype and refractive errors may reflect parallel molecular processes rather than a causal relationship, as the development of the SCN and circadian system is modulated by input from the eyes^52^. Various processes in the eye, including the length of the eye^5^, display a daily rhythm, which is abolished in animals developing myopia^6^. If we consider chronotype as a marker for circadian rhythms on a molecular level, it may suggest that genetic variations that lengthen the circadian clock and result in late chronotype may also lead to excessive eye elongation by disrupting the rhythmicity of eye growth. Further genetic studies and investigations in animal models are required to uncover the causality of this finding.

Intriguingly, we witnessed that decreased sleep duration and increased SJL increased the odds of myopia and hyperopia. SJL is indicative of the discrepancy between the inner biological clock of an individual and the social clock imposed by society^12^, and it has been associated with several adverse health outcomes, including metabolic disorders, cardiovascular problems, and psychiatric diseases^11^. It can be hypothesised that increased SJL may disrupt processes governing eye development. Alternatively, an increased SJL might be desirable—individuals generally accumulate sleep debt over workdays, which is counterbalanced by longer sleep on the weekends^54^. A larger difference between the mid-point of sleep on working and free days can thus indicate voluntary sleep extension on the weekends, which can serve to dissipate sleep debt^54^, complicating the interpretation of this finding.

Additionally, we provide evidence that some factors protective of myopia may be risk factors for hyperopia. We demonstrate that an increased amount of time in daylight, which is protective of myopia^31–33^, associates with higher odds of hyperopia. Additionally, a higher level of education, which is linked with myopia, decreased the odds of hyperopia. While it has been shown that children with uncorrected hyperopia have worse academic performance^55^, moderate hyperopia has previously not been associated with a lower level of education in adulthood^56^. These findings further suggest that myopia and hyperopia are conditions on the same spectrum, and specific factors affect the conditions in opposing directions. This insight is crucial for further analysis of the factors underlying myopia, highlighting several elements that predispose individuals to refractive errors in general, as well as those influencing their directionality.

In addition to investigating circadian rhythms and sleep in refractive errors, we characterise chronotype and sleep parameters in the EstBB. As reported previously, we found that chronotype decreased throughout adulthood, showed differences between sexes^12,36^, and SJL also decreased with age^10,12^. As a novel finding, we characterise the sex differences of SJL. Since late chronotype has been associated with increased SJL^12^ and men have a later chronotype until around the 50s^12,36^, men have also been hypothesised to display larger SJL^11^. Surprisingly, we found that SJL was larger in women from the 30s until late adulthood when SJL became negligible (Fig. 1b, Table 1). These findings indicate that although SJL is influenced by chronotype, these two measures are distinct and characterise specific aspects of the interaction between circadian rhythms and sleep.

While providing valuable insights into the relationship between refractive errors, chronotype and sleep, this study is subject to certain limitations that warrant consideration. First, although the sample is representative of the Estonian population^57^ and has sufficient statistical power, the ethnic diversity within the EstBB is relatively low (see Table 2). It is known that circadian rhythms are affected by ethnicity^58,59^, and it is, therefore, challenging to generalise our findings to non-European populations. Consequently, it is crucial to replicate the results presented here in more diverse and also non-European settings to ascertain their broader applicability and to understand the potential variances in these associations across different populations. Second, we defined myopia and hyperopia using data from electronic health records and self-reports, which do not indicate the extent of the refractive errors. It would be insightful to conduct similar analyses with refractive error as a continuous variable to determine whether the extreme chronotypes would also be associated with extreme refractive errors. Such studies would further strengthen the finding of an inverse relationship between chronotype and refractive errors.

To unravel this complex interplay between circadian rhythms and refractive errors, further research using a variety of approaches is required. Genetic studies, particularly those leveraging large biobank datasets, could be instrumental in identifying specific genes that contribute to this association. Such genetic insights would enhance our understanding of the molecular mechanisms of eye development and aid in developing targeted therapeutic interventions. Additionally, studies in animal models will be invaluable in dissecting causality and molecular mechanisms that link circadian rhythms with refractive development. Animal models enable the manipulation of genes, molecular pathways, and environmental conditions in a controlled fashion, allowing the detailed study of these interactions. Finally, there is a compelling need for additional observational studies, especially those focusing on critical developmental stages such as childhood and adolescence, to validate and extend the correlations identified in this work. Longitudinal studies tracking individuals over time would be particularly informative, as they could establish temporal relationships and potentially causal links between chronotype, sleep patterns, and refractive development. These investigations should also elucidate the interaction of environmental factors, such as light exposure and lifestyle habits, with genetic predispositions that could provide a more holistic understanding of refractive development. This could lead to more effective prevention strategies tailored to individual risk profiles, incorporating both genetic and environmental considerations.

## Methods

### Data in the Estonian Biobank

The Estonian Biobank (EstBB) is a population-based biobank with 210,500 adult participants in the 20^th^ December 2022 data freeze^57^, which was used for this study. All biobank participants have signed a broad informed consent form. Information on medical diagnoses is obtained through annual linking with the Estonian National Health Insurance Fund and other relevant databases. Diagnoses are coded with the International Classification of Disease 10^th^ edition (ICD-10). The majority of the electronic health records have been collected since 2004^57^. The activities of the EstBB are regulated by the Human Genes Research Act^60^, which was adopted in 2000 specifically for the operations of EstBB. Individual level data analysis in EstBB was carried out under ethical approval 1.1-12/624 from the Estonian Committee on Bioethics and Human Research (Estonian Ministry of Social Affairs), using data according to release application 3-10/GI/34223 from the EstBB.

All EstBB participants have been genotyped at the Core Genotyping Lab of the Institute of Genomics, University of Tartu, using Illumina Global Screening Array v3.0_EST. Samples were genotyped and PLINK format files were created using Illumina GenomeStudio v2.0.4. Individuals were excluded from the analysis if their SNP call-rate was <95%, if they were outliers of the absolute value of heterozygosity (>3 standard deviations from the mean) or if sex defined based on heterozygosity of X chromosome did not match sex in phenotype data^61^. Genotyped variant positions were in build GRCh37, and genetic principal components analysis (PCA) was performed in PLINK2^62^. Ancestry proportions and ancestry grouping were estimated with bigsnpr^63^. For ancestry inference, genotypes were imputed using 1000 Genome Project samples (n=2495). The total number of SNPs used for the inference was ∼4.54 million. Samples from all ancestries were included in the study.

### Munich Chronotype Questionnaire

The standard questionnaire administered to all EstBB participants contains the Estonian translation^64^ of the Munich Chronotype Questionnaire (MCTQ)^9^. The questionnaire asks participants about their sleep habits over the past four weeks separately for working and free (non-working) days. Participants were included in the analysis only if they reported not having done any shift work in the past three months and waking up without an alarm clock on free days, as only then is the calculated chronotype value estimating true biological clock^9^. All sleep parameters were calculated as per Roenneberg *et al*. 2003^9^. Briefly, chronotype was defined as the mid-point of sleep on free days adjusted for sleep debt (MSFsc) and reported as local time^9^ in 24-hour format. SJL was defined as the absolute difference between one’s midpoint of sleep on free and working days. Average daily sleep duration was calculated as (sleep duration on a working day * 5 + sleep duration on a free day * 2) / 7. Average daily exposure to daylight was calculated as (time in daylight on a working day * 5 + time in daylight on a free day * 2) / 7. Replies were filtered for unrealistic and extreme values (see Supplementary Fig. 1 for workflow). MCTQ parameters were characterised in participants ≤80 years at the time of replying to MCTQ.

### Definition of myopia and hyperopia

Participants were classified as myopes if they had received the ICD-10 diagnosis of H52.1 (myopia) or if they reported having received the diagnosis of myopia; and hyperopes if they had received the diagnosis of H52.0 (hyperopia) or if they reported having received the diagnosis of hyperopia. Self-reported diagnoses constituted 11% and 13% of the total number of diagnoses for myopia and hyperopia, respectively. Self-reported diagnoses were included as in Estonia, people with refractive errors are often followed primarily by optometrists, whose diagnoses are not reflected in the National Electronic Health records. Participants who had both myopia as well as hyperopia were excluded from the analysis (Supplementary Fig. 1). All remaining participants were defined as controls.

### Additional sociodemographic characteristics included in the analyses

The highest level of education was self-reported by the participants on a nine-level scale, and these were further combined into three categories as follows: 1) basic education (no primary education, primary education, and basic education); 2) secondary education (secondary education and vocational secondary education); 3) higher education (higher education, applied higher education and advanced academic degree). The education level reported at the time of filling out the MCTQ was used. Daily time spent in daylight was reported by participants as part of the MCTQ separately for weekdays and free days. The average daily time spent in daylight was calculated as follows: (time spent in daylight on a working day * 5 + time spent in daylight on a free day * 2) / 7. The season of birth was defined as follows. Spring: March, April, May, summer: June, July, August; autumn: September, October, November; winter: December, January, February. Photoperiod categories were defined as per Mandel *et al*. 2008^34^. Briefly, the civil twilight hours (period from dawn until dusk) in Tallinn in 2022 were downloaded from the US Navy Astronomical Applications Department public repository (available at: https://aa.usno.navy.mil/data/Dur_OneYear, accessed August 31, 2023). Tallinn was chosen as the reference as it is home to approx. 33% of the Estonian population. The 365 days of the year were divided into four photoperiods so that they would include approximately an equal number of days. Photoperiods were defined as shortest (6 hours 3 minutes to 8 hours 23 minutes of daylight; November 6 – February 1, 90 days), followed by short (8 hours 25 minutes to 12 hours 21 minutes of daylight; February 2 – March 21 and September 21 – Nov 5, 91 days), long photoperiod (12 hours 23 minutes to 16 hours 18 minutes of daylight; March 22 – May 5 and August 6 – September 20, 92 days) and longest photoperiod (16 hours 18 minutes to 18 hours and 39 minutes of daylight, May 6 – August 5, 92 days). Age at filling out the MCTQ was used as the age reported in the article. Age was included as a covariate in the logistic regression analyses with one-month precision. MCTQ parameters were characterised for participants aged ≤80 years to allow for comparison with previously published datasets^12,65^. Logistic regression analysis was performed in participants aged ≤70 years old as age-related diseases, e.g. ocular and neurological conditions and eye diseases, can influence one’s sleep patterns and circadian rhythms^41,42^.

### Statistical analysis

Descriptive statistics (mean and standard deviation; median and interquartile range; number and proportion) were used to characterise the study population and refractive error subgroups. Variables characterised included age, sex, education, birth season, photoperiod at birth, ethnicity, chronotype, SJL, sleep duration (SDweek) and daylight exposure.

Chronotype, SJL and average sleep duration in a day were characterised by sex across age groups. The study population was divided into 12 age groups, the youngest aged 18-25 years, followed by 5-year intervals up to age 80. For variables with normal distribution (MSFsc and SDweek), a linear regression model including an interaction term between sex and age group was fitted. The emmeans R package version 1.8.9^66^ was used further to compare MSFsc and SDweek between sexes within each age group and to estimate marginal means with 95% confidence intervals. Differences between sexes were analysed with two-tailed t-tests followed by the Bonferroni method to control for the family-wise error rate. To estimate medians and 95% confidence intervals of medians of SJL across sexes and age groups, bootstrapping using the boot R package version 1.3-28.1^67^ was employed. A quantile regression model was fitted using the quantreg R package version 5.96^68^ including an interaction term between sex and age group. Differences in median values between the sexes within each age group were assessed using the *p* values associated with the interaction terms between sex and each age group in the quantile regression model. Correlations between chronotype, SJL, average sleep duration and time spent in daylight were estimated using Pearson and Spearman correlation for normally and non-normally distributed variables, respectively.

To assess the association between a set of predictors and myopia/hyperopia, we fitted separate logistic regression models with myopia or hyperopia status as the dependent variable, including participants classified as myopes or hyperopes as cases, respectively and participants with no refractive error as controls. For univariate analysis, a separate model adjusted for age, age squared, and sex was fitted for each predictor (chronotype, SJL, average sleep duration, time spent in daylight, education level, season at birth and photoperiod at birth). For multivariate analyses, all the aforementioned predictors were simultaneously analysed, and the models were additionally adjusted for the first ten genetic principal components. Multicollinearity was tested by calculating the variance inflation factor for independent variables in regression models, and no multicollinearity was detected between the variables of interest. As a sensitivity analysis, the same multivariate analyses were pursued in a subsample with age >45 years. All analyses were carried out and visualised using R 4.3.1^69^ and RStudio 2023.06.1+524^70^. A *p* value of <0.05 was considered statistically significant.

### Inclusion & Ethics

All biobank participants have signed a broad informed consent form, and information on ICD-10 diagnosis codes is obtained via regular linking with the National Health Insurance Fund and other relevant databases, with the majority of the electronic health records having been collected since 2004^57^. The activities of the EstBB are regulated by the Human Genes Research Act^60^, which was adopted in 2000 specifically for the operations of EstBB. Individual level data analysis in EstBB was carried out under ethical approval 1.1-12/624 from the Estonian Committee on Bioethics and Human Research (Estonian Ministry of Social Affairs), using data according to release application 3-10/GI/34223 from the EstBB.

## Acknowledgements

This research was supported by the European Union through Horizon 2020 research and innovation programme under grants no 810645 and 894987, through the European Regional Development Fund project no. MOBEC008, and the Estonian Research Council grants PRG1291 and PRG1414. Data analysis was carried out in part in the High-Performance Computing Centre of the University of Tartu. We acknowledge the Estonian Biobank Research Team (Andres Metspalu, Reedik Mägi, Mari Nelis and Georgi Hudjashov). We would like to thank Priit Palta for constructive feedback on the manuscript.

## Author contributions

T.P. and E.A. designed the study, T.P., M.T.-L. and K.-L.K. performed the MCTQ calculations and data cleaning. T.P and N.T. performed the sample characterisation and statistical analyses. U.A., E.A. and K.R conducted the genetic analyses for genetic principal components and ancestry. E.A and T.E supervised the study. T.P. and E.A. wrote the first draft of the manuscript. All authors critically reviewed the manuscript. E.A. and T.E. contributed equally to this work.

## Data availability

Pseudonymised genotype and phenotype data are available from the Estonian Biobank (https://genomics.ut.ee/en/content/estonian-biobank) upon request.

## Code availability

Analyses used are described in the Methods section of the manuscript.

## References

1. Holden, B. A. et al. Global Prevalence of Myopia and High Myopia and Temporal Trends from 2000 through 2050. Ophthalmology 123, 1036–1042 (2016).

2. Flitcroft, D. I. Emmetropisation and the aetiology of refractive errors. Eye (Lond) 28, 169– 179 (2014).

3. Tideman, J. W. L. et al. Association of Axial Length With Risk of Uncorrectable Visual Impairment for Europeans With Myopia. JAMA Ophthalmol 134, 1355–1363 (2016).

4. Holy, C., Kulkarni, K. & Brennan, N. A. Predicting Costs and Disability from the Myopia Epidemic – A Worldwide Economic and Social Model. Investig Ophthalmol Vis Sci 60, 5466 (2019).

5. Chakraborty, R., Read, S. & Collins, M. Diurnal Variations in Axial Length, Choroidal Thickness, Intraocular Pressure, and Ocular Biometrics. Invest Ophthalmol Vis Sci 52, 5121–5129 (2011).

6. Nickla, D. L., Wildsoet, C. & Wallman, J. Visual Influences on Diurnal Rhythms in Ocular Length and Choroidal Thickness in Chick Eyes. Exp Eye Res 66, 163–181 (1998).

7. Weiss, S. & Schaeffel, F. Diurnal growth rhythms in the chicken eye: relation to myopia development and retinal dopamine levels. J Comp Physiol A 172, 263–270 (1993).

8. Hysi, P. G. et al. Meta-analysis of 542,934 subjects of European ancestry identifies new genes and mechanisms predisposing to refractive error and myopia. Nat Genet 52, 401– 407 (2020).

9. Roenneberg, T., Wirz-Justice, A. & Merrow, M. Life between clocks: daily temporal patterns of human chronotypes. J Biol Rhythms 18, 80–90 (2003).

10. Wittmann, M., Dinich, J., Merrow, M. & Roenneberg, T. Social Jetlag: Misalignment of Biological and Social Time. Chronobiology International 23, 497–509 (2006).

11. Caliandro, R., Streng, A. A., van Kerkhof, L. W. M., van der Horst, G. T. J. & Chaves, I. Social Jetlag and Related Risks for Human Health: A Timely Review. Nutrients 13, 4543 (2021).

12. Roenneberg, T. et al. A marker for the end of adolescence. Current Biology 14, R1038– R1039 (2004).

13. Sun, J. et al. Chronotype: implications for sleep quality in medical students. Chronobiol Int 36, 1115–1123 (2019).

14. Rique, G. L. N., Fernandes Filho, G. M. C., Ferreira, A. D. C. & de Sousa-Muñoz, R. L. Relationship between chronotype and quality of sleep in medical students at the Federal University of Paraiba, Brazil. Sleep Sci 7, 96–102 (2014).

15. Lima, Á. M. A., Varela, G. C. G., Silveira, H. A. da C., Parente, R. D. G. & Araujo, J. F. Evening chronotypes experience poor sleep quality when taking classes with early starting times. Sleep Sci 3, 45–48 (2010).

16. Ayaki, M., Torii, H., Tsubota, K. & Negishi, K. Decreased sleep quality in high myopia children. Sci Rep 6, 33902–33902 (2016).

17. Liu, X. N. et al. Sleeping late is a risk factor for myopia development amongst school-aged children in China. Sci Rep 10, 17194 (2020).

18. Lee, S. S.-Y. & Mackey, D. A. Prevalence and Risk Factors of Myopia in Young Adults: Review of Findings From the Raine Study. Front. Public Health 10, 861044 (2022).

19. Xu, S. et al. Association between sleep-wake schedules and myopia among Chinese school-aged children and adolescents: a cross-sectional study. BMC Ophthalmol 23, 135 (2023).

20. Wei, S.-F. et al. Sleep Duration, Bedtime, and Myopia Progression in a 4-Year Follow-up of Chinese Children: The Anyang Childhood Eye Study. Invest. Ophthalmol. Vis. Sci. 61, 37–37 (2020).

21. Ostrin, L. A., Read, S. A., Vincent, S. J. & Collins, M. J. Sleep in Myopic and Non-Myopic Children. Trans. Vis. Sci. Tech. 9, 22–22 (2020).

22. Li, M. et al. Sleep Patterns and Myopia Among School-Aged Children in Singapore. Front Public Health 10, 828298 (2022).

23. Peng, W., Sun, S. M., Wang, F. & Sun, Y. N. Comparison of Factors Associated with Myopia among Middle School Students in Urban and Rural Regions of Anhui, China. Optometry and Vision Science 99, 702 (2022).

24. Zhou, Z. et al. Disordered Sleep and Myopia Risk among Chinese Children. PLoS One 10, e0121796 (2015).

25. Stafford-Bell, N. et al. Associations of 12-year sleep behaviour trajectories from childhood to adolescence with myopia and ocular biometry during young adulthood. Ophthalmic Physiol Opt 42, 19–27 (2022).

26. Jee, D., Morgan, I. G. & Kim, E. C. Inverse relationship between sleep duration and myopia. Acta Ophthalmol 94, e204–210 (2016).

27. Chakraborty, R. et al. Myopia, or near-sightedness, is associated with delayed melatonin circadian timing and lower melatonin output in young adult humans. Sleep 44, zsaa208 (2021).

28. Chakraborty, R. et al. Delayed melatonin circadian timing, lower melatonin output, and sleep disruptions in myopic, or short-sighted, children. Sleep 47, zsad265 (2024).

29. Flanagan, S. C., Cobice, D., Richardson, P., Sittlington, J. J. & Saunders, K. J. Elevated Melatonin Levels Found in Young Myopic Adults Are Not Attributable to a Shift in Circadian Phase. Invest. Ophthalmol. Vis. Sci. 61, 45–45 (2020).

30. Pärssinen, O. The increased prevalence of myopia in Finland. Acta Ophthalmologica 90, 497–502 (2012).

31. Rose, K. A. et al. Outdoor Activity Reduces the Prevalence of Myopia in Children. Ophthalmology 115, 1279–1285 (2008).

32. Xiong, S. et al. Time spent in outdoor activities in relation to myopia prevention and control: a meta-analysis and systematic review. Acta Ophthalmol 95, 551–566 (2017).

33. Ho, C.-L., Wu, W.-F. & Liou, Y. M. Dose–Response Relationship of Outdoor Exposure and Myopia Indicators: A Systematic Review and Meta-Analysis of Various Research Methods. IJERPH 16, 2595 (2019).

34. Mandel, Y. et al. Season of Birth, Natural Light, and Myopia. Ophthalmology 115, 686– 692 (2008).

35. McMahon, G. et al. Season of Birth, Daylight Hours at Birth, and High Myopia. Ophthalmology 116, 468–473 (2009).

36. Fischer, D., Lombardi, D. A., Marucci-Wellman, H. & Roenneberg, T. Chronotypes in the US – Influence of age and sex. PLOS ONE 12, e0178782 (2017).

37. Campbell, S. S. & Murphy, P. J. The nature of spontaneous sleep across adulthood. J Sleep Res 16, 24–32 (2007).

38. Dorffner, G., Vitr, M. & Anderer, P. The effects of aging on sleep architecture in healthy subjects. Adv Exp Med Biol 821, 93–100 (2015).

39. Ohayon, M. M., Carskadon, M. A., Guilleminault, C. & Vitiello, M. V. Meta-Analysis of Quantitative Sleep Parameters From Childhood to Old Age in Healthy Individuals: Developing Normative Sleep Values Across the Human Lifespan. Sleep 27, 1255–1273 (2004).

40. Boubekri, M., Cheung, I. N., Reid, K. J., Wang, C.-H. & Zee, P. C. Impact of Windows and Daylight Exposure on Overall Health and Sleep Quality of Office Workers: A Case-Control Pilot Study. Journal of Clinical Sleep Medicine 10, 603–611.

41. Lee, S. S. Y., Nilagiri, V. K. & Mackey, D. A. Sleep and eye disease: A review. Clinical & Experimental Ophthalmology 50, 334–344 (2022).

42. Musiek, E. S. & Holtzman, D. M. Mechanisms linking circadian clocks, sleep, and neurodegeneration. Science 354, 1004–1008 (2016).

43. Williams, K. M. et al. Prevalence of refractive error in Europe: the European Eye Epidemiology (E3) Consortium. Eur J Epidemiol 30, 305–315 (2015).

44. Lane, J. M. et al. Genome-wide association analysis identifies novel loci for chronotype in 100,420 individuals from the UK Biobank. Nat Commun 7, 10889 (2016).

45. Jones, S. E. et al. Genome-Wide Association Analyses in 128,266 Individuals Identifies New Morningness and Sleep Duration Loci. PLOS Genetics 12, e1006125 (2016).

46. Jones, S. E. et al. Genome-wide association analyses of chronotype in 697,828 individuals provides insights into circadian rhythms. Nat Commun 10, 343 (2019).

47. Vink, J. M., Vink, J. M., Groot, A. S., Kerkhof, G. A. & Boomsma, D. I. Genetic Analysis of Morningness and Eveningness. Chronobiology International 18, 809–822 (2001).

48. Jones, C. R. et al. Familial advanced sleep-phase syndrome: A short-period circadian rhythm variant in humans. Nat Med 5, 1062–1065 (1999).

49. Toh, K. L. et al. An hPer2 Phosphorylation Site Mutation in Familial Advanced Sleep Phase Syndrome. Science 291, 1040–1043 (2001).

50. Xu, Y. et al. Functional consequences of a CKIδ mutation causing familial advanced sleep phase syndrome. Nature 434, 640–644 (2005).

51. Schamilow, S. et al. Time Spent Outdoors and Associations with Sleep, Optimism, Happiness and Health before and during the COVID-19 Pandemic in Austria. Clocks Sleep 5, 358–372 (2023).

52. Wong, S. D. et al. Development of the circadian system in early life: maternal and environmental factors. Journal of Physiological Anthropology 41, 22 (2022).

53. Poudel, S. et al. Contrast Sensitivity of ON and OFF Human Retinal Pathways in Myopia. J. Neurosci. 44, (2024).

54. Roenneberg, T. How can social jetlag affect health? Nat Rev Endocrinol 19, 383–384 (2023).

55. Mavi, S. et al. The Impact of Hyperopia on Academic Performance Among Children: A Systematic Review. Asia-Pacific Journal of Ophthalmology 11, 36–51 (2022).

56. Plotnikov, D., Sheehan, N. A., Williams, C., Atan, D. & Guggenheim, J. A. Hyperopia Is Not Causally Associated With a Major Deficit in Educational Attainment. Transl Vis Sci Technol 10, 34 (2021).

57. Leitsalu, L. et al. Cohort Profile: Estonian Biobank of the Estonian Genome Center, University of Tartu. International Journal of Epidemiology 44, 1137–1147 (2015).

58. Eastman, C. I., Suh, C., Tomaka, V. A. & Crowley, S. J. Circadian rhythm phase shifts and endogenous free-running circadian period differ between African-Americans and European-Americans. Sci Rep 5, 8381 (2015).

59. Malone, S. K., Patterson, F., Lu, Y., Lozano, A. & Hanlon, A. Ethnic differences in sleep duration and morning–evening type in a population sample. Chronobiology International 33, 10–21 (2016).

60. Riigikogu Human Genes Research Act. (2000).

61. Mitt, M. et al. Improved imputation accuracy of rare and low-frequency variants using population-specific high-coverage WGS-based imputation reference panel. Eur J Hum Genet 25, 869–876 (2017).

62. Chang, C. C. et al. Second-generation PLINK: rising to the challenge of larger and richer datasets. GigaScience 4, s13742-015-0047–8 (2015).

63. Privé, F., Aschard, H., Ziyatdinov, A. & Blum, M. G. B. Efficient analysis of large-scale genome-wide data with two R packages: bigstatsr and bigsnpr. Bioinformatics 34, 2781– 2787 (2018).

64. Allebrandt, K. V. et al. CLOCK gene variants associate with sleep duration in two independent populations. Biol Psychiatry 67, 1040–1047 (2010).

65. Roenneberg, T., Allebrandt, K. V., Merrow, M. & Vetter, C. Social Jetlag and Obesity. Current Biology 22, 939–943 (2012).

66. Lenth, R. V. emmeans: Estimated Marginal Means, aka Least-Squares Means. (2023).

67. Canty, A. & Ripley, B. D. boot: Bootstrap R (S-Plus) Functions. (2022).

68. Koenker, R. quantreg: Quantile Regression. (2023).

69. R Core Team. R: A language and environment for statistical computing. R Foundation for Statistical Computing (2023).

70. RStudio Team. RStudio: Integrated Development Environment for R. RStudio, PBC. (2020).

