## Supplementary Information for "Myopia and hyperopia are associated with opposite chronotypes in a sample of 71,016 individuals"

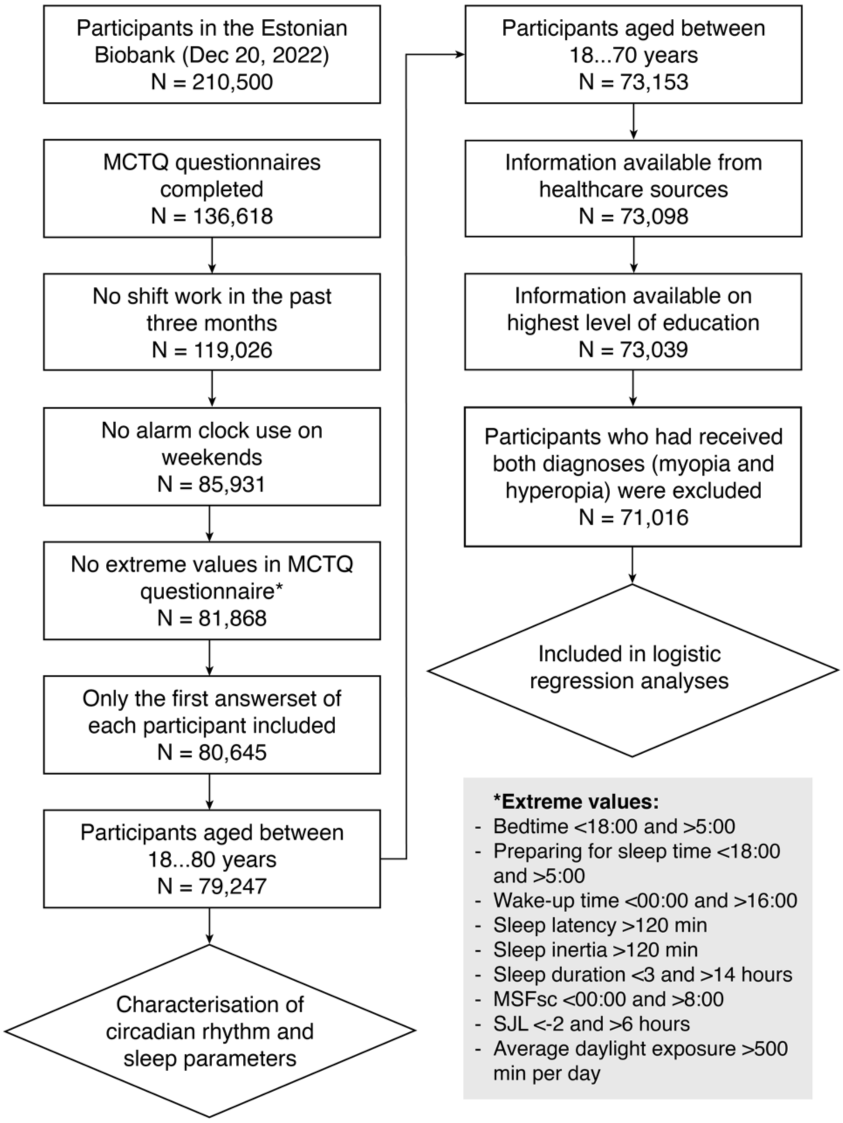


**Supplementary Figure 1. Flow diagram of study sample selection.**
Sleep latency is defined in the MCTQ as the self-reported amount of time it takes to fall asleep. Sleep inertia is the self-reported time it takes to get up after waking up.
MCTQ –Munich Chronotype Questionnaire, MSFsc – mid-point of sleep on free days adjusted for sleep debt, SJL – social jet lag.

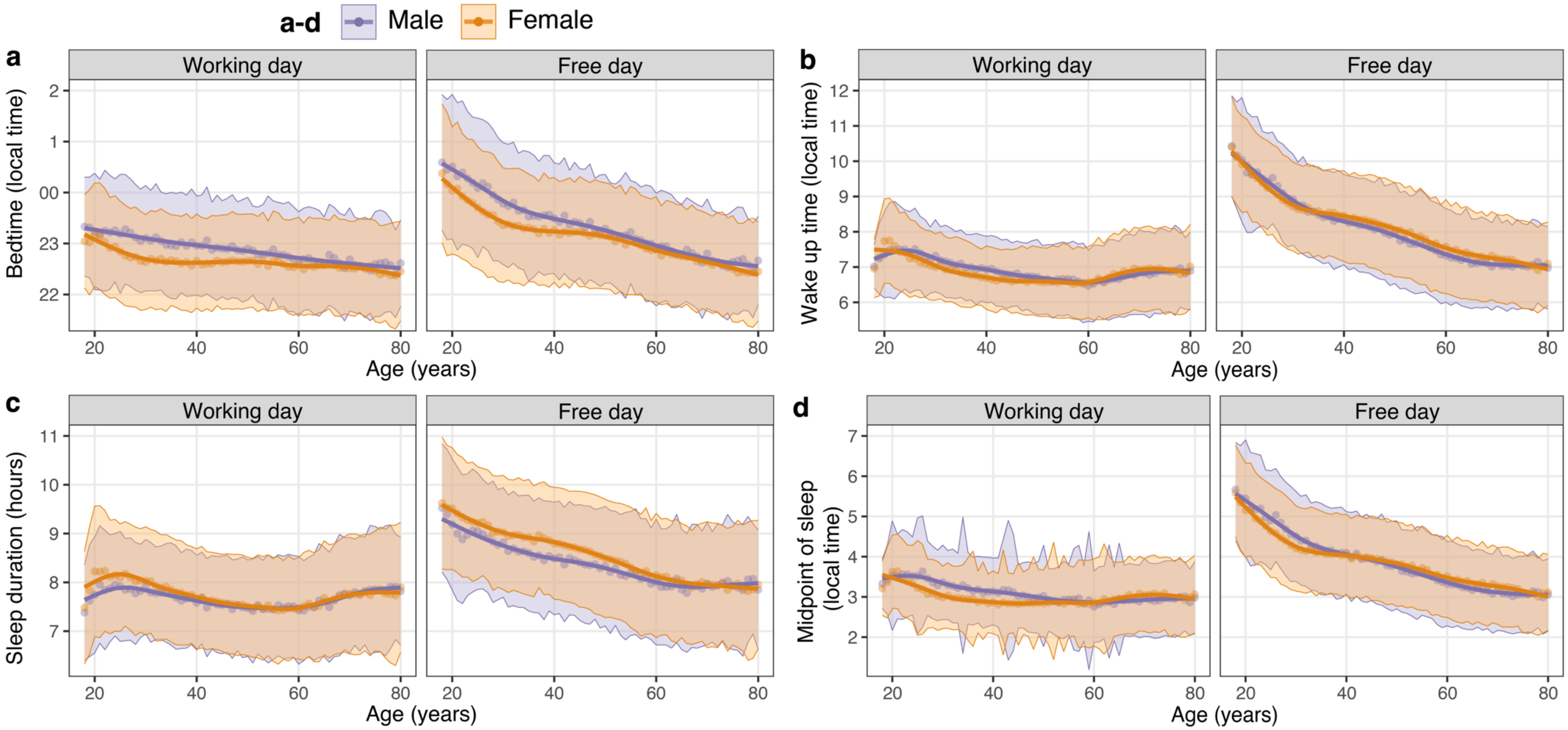


**Supplementary Figure 2. Characterisation of sleep timing on working and free days in the Estonian Biobank.**
**a-d** Local time of going to bed **(a)** and waking up **(b)**, sleep duration **(c)**, and midpoint of sleep **(d)** for working and free days across ages in one-year bins in males and females. Points display means for sex and age, highlighted areas indicate ±SD and means are fitted with a generalised additive model. Only participants who reported waking up without an alarm clock and no shift work three months prior to replying were included.

**Supplementary Table 1. Association of myopia and hyperopia with sleep and circadian rhythm parameters in univariate analyses**

| | Myopia | | | Hyperopia | | |
|---|---|---|---|---|---|---|
| **Predictors** | **Odds Ratio** | **95% CI** | ***p* value** | **Odds Ratio** | **95% CI** | ***p* value** |
| Chronotype (MSFsc, per hour) | 1.07 | 1.05 – 1.09 | **<0.001** | 0.94 | 0.92 – 0.96 | **<0.001** |
| SJL (per hour) | 1.06 | 1.04 – 1.09 | **<0.001** | 1.05 | 1.02 – 1.08 | **0.003** |
| SDweek (per hour) | 0.95 | 0.93 – 0.97 | **<0.001** | 0.91 | 0.89 – 0.93 | **<0.001** |
| LEweek (per hour) | 0.93 | 0.92 – 0.95 | **<0.001** | 1.07 | 1.06 – 1.09 | **<0.001** |
| Education | | | | | | |
| Middle | *Reference* | | | *Reference* | | |
| Basic | 0.60 | 0.56 – 0.65 | **<0.001** | 1.02 | 0.94 – 1.11 | 0.629 |
| Higher | 1.44 | 1.38 – 1.49 | **<0.001** | 0.69 | 0.66 – 0.73 | **<0.001** |
| Birth season | | | | | | |
| Winter | *Reference* | | | *Reference* | | |
| Spring | 1.02 | 0.97 – 1.07 | 0.41 | 1.06 | 0.99 – 1.13 | 0.10 |
| Summer | 1.02 | 0.97 – 1.07 | 0.45 | 1.04 | 0.98 – 1.11 | 0.20 |
| Autumn | 1.04 | 0.99 – 1.10 | 0.08 | 1.07 | 1.00 – 1.14 | **0.04** |
| Photoperiod at birth (hh:mm) | | | | | | |
| Shortest (06:03-08:23) | *Reference* | | | *Reference* | | |
| Short (08:25-12:21) | 0.98 | 0.93 – 1.03 | 0.39 | 1.06 | 0.99 – 1.13 | 0.11 |
| Long (12:23-16:18) | 1.00 | 0.95 – 1.05 | 0.85 | 1.04 | 0.98 – 1.11 | 0.22 |
| Longest (16:18-18:39) | 1.03 | 0.98 – 1.08 | 0.30 | 1.07 | 1.01 – 1.15 | **0.031** |
| Observations | 60,348 | | | 52,500 | | |

Odds ratios were obtained from univariate logistic regression models with myopia or hyperopia status as the dependent variable, including participants classified as myopes or hyperopes as cases, respectively, and participants with no refractive error as controls. Separate models were fitted for each predictor, while adjusting for age, age squared and sex. *p* values < 0.05 are highlighted in bold.
LEweek –average daily exposure to daylight, MSFsc – mid-point of sleep on free days adjusted for sleep debt, SDweek – weekly average sleep duration, SJL – social jet lag, CI – confidence interval.

**Supplementary Table 2. Association of myopia and hyperopia with sleep and circadian rhythm parameters in individuals aged >45 years**

| | Myopia | | | Hyperopia | | |
|---|---|---|---|---|---|---|
| **Predictors** | **Odds Ratio** | **95% CI** | ***p* value** | **Odds Ratio** | **95% CI** | ***p* value** |
| Chronotype (MSFsc, per hour) | 1.04 | 1.01 – 1.07 | **0.023** | 0.93 | 0.91 – 0.96 | **<0.001** |
| SJL (per hour) | 1.04 | 1.00 – 1.09 | **0.047** | 1.08 | 1.03 – 1.12 | **0.001** |
| SDweek (per hour) | 0.95 | 0.92 – 0.98 | **0.001** | 0.91 | 0.89 – 0.94 | **<0.001** |
| LEweek (per hour) | 0.97 | 0.95 – 0.99 | **0.002** | 1.06 | 1.04 – 1.08 | **<0.001** |
| Education | | | | | | |
| Middle | *Reference* | | | *Reference* | | |
| Basic | 0.60 | 0.51 – 0.70 | **<0.001** | 0.98 | 0.88 – 1.09 | 0.70 |
| Higher | 1.43 | 1.35 – 1.52 | **<0.001** | 0.77 | 0.72 – 0.81 | **<0.001** |
| Birth season | | | | | | |
| Winter | *Reference* | | | *Reference* | | |
| Spring | 1.12 | 0.99 – 1.27 | 0.07 | 1.06 | 0.94 – 1.19 | 0.37 |
| Summer | 1.13 | 0.98 – 1.30 | 0.10 | 1.02 | 0.89 – 1.17 | 0.73 |
| Autumn | 1.06 | 0.97 – 1.16 | 0.19 | 1.04 | 0.95 – 1.13 | 0.39 |
| Photoperiod at birth (hh:mm) | | | | | | |
| Shortest (06:03-08:23) | *Reference* | | | *Reference* | | |
| Short (08:25-12:21) | 0.87 | 0.80 – 0.95 | **0.002** | 1.03 | 0.95 – 1.13 | 0.48 |
| Long (12:23-16:18) | 0.84 | 0.75 – 0.96 | **0.007** | 0.98 | 0.87 – 1.11 | 0.79 |
| Longest (16:18-18:39) | 0.85 | 0.74 – 0.98 | **0.026** | 1.05 | 0.92 – 1.21 | 0.46 |
| Sex, reference: male | 1.95 | 1.83 – 2.09 | **<0.001** | 1.76 | 1.66 – 1.87 | **<0.001** |
| Age | 1.05 | 0.98 – 1.12 | 0.20 | 1.36 | 1.27 – 1.46 | **<0.001** |
| Age squared /1,000 | 0.68 | 0.36 – 1.26 | 0.22 | 0.12 | 0.07 – 0.22 | **<0.001** |
| (Intercept) | 3.19 | 2.40 – 4.25 | **<0.001** | 0.18 | 0.12 – 0.26 | **<0.001** |
| Observations | 25,465 | | | 26,734 | | |

Odds ratios were obtained from two multivariable logistic regression models with myopia or hyperopia status as the dependent variable, including participants classified as myopes or hyperopes as cases, respectively, and participants with no refractive error as controls. Both models used MSFsc, SJL, SDweek, LEweek, education, birth season, and photoperiod at birth as predictors, and were adjusted for age, age squared, sex and first ten genetic principal components. *p* values < 0.05 are highlighted in bold.
LEweek – average daily exposure to daylight, MSFsc – mid-point of sleep on free days adjusted for sleep debt, SDweek – weekly average sleep duration, SJL – social jet lag, CI – confidence interval.
